# Chronic Low Back Pain in Young Adults: Pathophysiological Aspects of Neuroinflammation and Degeneration

**DOI:** 10.1101/2025.05.17.25327838

**Authors:** Natalya G. Pravdyuk, Anastasiia A. Buianova, Anna V. Novikova, Alesya A. Klimenko, Mikhail A. Ignatyuk, Liubov A. Malykhina, Olga I. Patsap, Dmitrii A. Atiakshin, Vitaliy V. Timofeev, Nadezhda A. Shostak

## Abstract

**Introduction:** Degenerative disc disease (DDD) is a major cause of lower back pain (LBP). Key pathological processes of intervertebral disc (IVD) degeneration include extracellular matrix (ECM) degradation (including aggrecan loss), cartilage dehydration, and pathological ingrowth of blood vessels and nerve fibers. Neurotrophins and neuropeptides, such as nerve growth factor (NGF) and substance P (SP), play an essential role in LBP pathogenesis, neoinnervation, inflammation, and the maintenance of chronic pain.

**Materials and Methods:** Thirty-six young patients (mean age 36.00 [31.00, 42.50] years) with LBP associated with herniated discs and five healthy individuals were enrolled. IVD samples were collected during microdiscectomy. MRI-based Pfirrmann classification (2001) was used to assess the stages of disc degeneration. Histological grading was performed according to Sive’s criteria (2002). Histochemical staining (hematoxylin-eosin, Alizarin Red, Safranin O/Fast Green FCF) was conducted to evaluate ECM status, including aggrecan content. Immunohistochemical analysis was performed to assess NGF, S-100 protein, and SP expression.

**Results:** All patients experienced chronic LBP. According to MRI, Pfirrmann grade V degeneration was found in 30.55% of patients, grade IV in 61.11%, grade III in 5.56%, and grade II in 2.78%. Histologically confirmed degeneration was observed in 23 cases (63.88%), with 3 patients showing severe degeneration (10-12 points). In patients with longer pain episodes (average duration 11.29 weeks), aggrecan loss was observed in 19.4% of cases (r=0.449; P=0.031). NGF expression was significantly higher in degenerated discs (P=0.0287) and positively correlated with SP levels (r=0.785; P=5.268 × 10^−9^). Increased NGF and SP expression were noted in patients with osteophytes, with levels correlating with both the histological degeneration score and MRI grading. Isolated free nerve endings were detected in the nucleus pulposus of 5 patients. Calcification was observed in 36.1% of cases, predominantly around hypertrophic chondrocytes and their clusters, and its severity correlated with radiculopathy (r=0.664; P=0.005).

**Conclusion:** In young individuals, aggrecan loss, increased expression of NGF, SP, S-100 protein, and ECM calcification are key pathological features of IVD degeneration contributing to chronic LBP. The colocalization of NGF and SP suggests a synergistic role in the development of chronic pain. These findings highlight new therapeutic targets aimed at inhibiting pathological neoinnervation and ECM degradation.

## Introduction

Low back pain (LBP) is one of the leading causes of disability and work incapacity worldwide, affecting individuals across all age groups [1, 2]. Increasingly, young adults are being affected, making LBP a growing public health concern. By 2050, the global burden of LBP is projected to rise by 36% [3, 4]. Among the various etiologies of LBP, over half are attributed to intervertebral disc (IVD) pathology [5].

The clinical syndrome characterized by back pain along with imaging-confirmed signs of disc damage is termed degenerative disc disease (DDD) [6]. DDD is primarily marked by disc dehydration, degradation of the extracellular matrix (ECM), formation of macroscopic cartilage defects, varying degrees of disc extrusion, neurovascular ingrowth, and sometimes calcification.

An IVD comprises three distinct components: the highly hydrated, proteoglycan-rich nucleus pulposus (NP); the annulus fibrosus (AF); and the cartilaginous endplates (EPs), which anchor the disc to the adjacent vertebral bodies [7]. In healthy individuals, NP hydration ranges between 70-90%, with its structural matrix consisting of 35-65% proteoglycans, 5-20% type II collagen, and small amounts of elastin and other proteins [8]. The NP contains large notochordal- derived cells and mesenchymal cells with chondrocyte-like morphology, which, during maturation and degeneration, transition into fibrochondrocytes [9, 10]. The AF consists of approximately 65-70% water, 20% proteoglycans, 50-70% type I collagen, and 2% elastin. Functionally, the IVD serves to support axial loads, absorb shock, allow spinal flexibility, and maintain intervertebral stability.

ECM degradation is manifested by the loss of structural macromolecules and hyaluronic acid components – most notably aggrecan, a key proteoglycan found in cartilage-rich disc tissue [11].

The IVD is an avascular structure. Its nutrition is maintained via capillaries supplying the peripheral AF [12] and through diffusion and osmosis across the cartilaginous EPs [13]. Under physiological conditions, only the outer layers of the AF are innervated by sinuvertebral nerves and branches from the ventral spinal roots and gray rami communicantes. These nerves comprise both small and large fibers functioning as mechanoreceptors and nociceptors [14].

During disc degeneration, infiltrating immune cells release pro-inflammatory cytokines, such as tumor necrosis factor-alpha (TNF-α), interleukins IL-1β, IL-6, and IL-17 [15, 16], which drive the degenerative cascade [17]. The synergistic role of these cytokines and neurotrophic factors – such as 1B4, calcitonin gene-related peptide (CGRP), nerve growth factor (NGF), brain-derived neurotrophic factor (BDNF), and neuropeptides like Substance P (SP), along with S-100 proteins – has been implicated in pain signal generation [18, 19]. In NP cells, even minimal concentrations of IL-1β (1 pg/mL) and NGF (≥0.1 ng/mL) significantly upregulate SP and NGF expression [19]. SP contributes to neovascularization within the degenerating IVD [20]. Chronic exposure to SP enhances pro-inflammatory cytokine production via NF-κB pathway activation, thereby promoting disc degeneration and discogenic pain [21]. It has been hypothesized that SP-positive nerve fibers in degenerated discs may be associated with neoangiogenesis [22].

S-100 proteins represent a group of calcium-binding, acidic peptides specific to neural tissue, exhibiting molecular and charge heterogeneity but immunological similarity [23]. Approximately 85-90% of total S-100 protein in the nervous system is localized in astrocytes, 10-15% in neurons, and trace amounts in oligodendrocytes. These proteins are synthesized by glial cells and subsequently transported to neurons. Intracellularly, S-100 proteins are primarily localized in the cytoplasm, synaptic membranes, and chromatin. Their main functions include regulation of intracellular processes (e.g., proliferation, apoptosis, inflammation, migration) and extracellular signaling (e.g., immune responses, tissue repair, invasion) through receptor-mediated mechanisms.

In the context of IVD degeneration associated with LBP, the roles of NGF and other neurotrophic markers are of increasing interest for understanding the molecular and functional interrelations across all disc components.

The objective of our study was to evaluate molecular and cellular markers of neo-innervation and ECM degradation in the IVD using immunohistochemical analysis in young adult patients with LBP.

## Materials and Methods

### Patient Samples

This study was conducted at the Pirogov Russian National Research Medical University between 2023 and 2024 and was approved by the local ethics committee (protocol No. 208, dated 17.05.2021). Informed consent was obtained from all participants. A total of 36 patients with chronic LBP and IVD herniation requiring surgical intervention were enrolled. The median age was Me = 36.00 [31.00, 42.50] years; 36.11% were female and 63.89% male. The average intensity of back pain on a visual analog scale was 71 [61.00, 90.00] mm. In 87% of the cases, the pain had a chronic course with duration ranging from 2 to 10 years (Table 1). Patients with infectious, traumatic, oncological, or rheumatic spinal lesions were excluded from the study.

**Table 1.** Clinical characteristics of patients based on instrumental diagnostic data (n = 36).

| Parameters | Values |
| --- | --- |
| Male | 23 (63.89%) |
| Female | 13 (36.11%) |
| Intensity of LBP on a visual analogue scale, mm | 71.00 [61.00, 90.00] |
| Duration of the latest pain episode, weeks | 4.00 [3.00, 10.00] |
| Presence of LBP | 36 (100 %) |
| Duration of chronic LBP, years | 5.00 [2.00, 10.00] |
| Number of exacerbations over the past 12 months | 2.00 [1.00, 2.00] |
| Stage of DDD according to Pfirrmann classification<br>• L1-L2<br>• L2-L3<br>• L3-L4<br>• L4-L5<br>• L5-S1 | 2.00 [1.00, 2.00]<br>2.00 [1.00, 2.00]<br>2.00 [1.00, 3.00]<br>4.00 [3.00, 4.00]<br>4.00 [4.00, 4.75] |
| Pfirrmann grade of the operated disc level, n (%)<br>• 2<br>• 3<br>• 4<br>• 5 | 1 (2.78%)<br>2 (5.56%)<br>22 (61.11%)<br>11 (30.55%) |
| Level of disc herniation, n (%)<br>• L4-L5, L5-S1<br>• L4-L5<br>• L5-S1 | 12 (33.33%)<br>4 (11.11%)<br>19 (52.78%) |
| Erosive changes of the EPs | 21 (58.33%) |
| Marginal osteophytes of the vertebral bodies | 16 (66.67%) |
| Facet joint osteoarthritis | 33 (91.67%) |
| Modic changes (Types I and II), n (%)<br>• Type I<br>• Type II | 20 (56.5%)<br>12 (60%)<br>8 (40%) |

Notes: LBP – low back pain; NP – nucleus pulposus; T2WI – T2-weighted imaging; EPs – vertebral endplates adjacent to the intervertebral disc at the level of herniation.

According to magnetic resonance imaging (MRI) of the lumbar spine, the stage of disc degeneration at the level of extrusion ranged from grade III to grade V according to the Pfirrmann classification, and was categorized as advanced (grade III), severe (grade IV), and irreversible (grade V), due to complete disorganization of the NP and significant dehydration. MRI revealed erosive endplate damage in 58.33% of cases, marginal vertebral osteophytes in 66.67%, and reactive changes in the adjacent vertebral bodies (Modic changes) in 56.52% of patients (60% Type I, 40% Type II). Facet joint osteoarthritis was present in 91.67% of cases.

Disc tissue samples from the study group were obtained during discectomy. Control samples were collected from five previously healthy individuals (median age Me = 40.00 [38.50, 42.00] years, all male) who had sustained acute motor vehicle trauma and underwent spinal stabilization surgery. Intervertebral cartilage segments (L4–L5, L5–S1) were excised within one hour post-trauma. In all biopsy samples, the NP was present and served as the primary object of investigation.

### Histochemical Staining

To identify degenerative changes in addition to the Pfirrmann-based scale, we employed hematoxylin and eosin (H&E), alizarin red, and trichrome staining.

Immediately following discectomy, IVD samples were fixed in 10% buffered formalin (pH = 7.4). The samples were embedded in paraffin, sectioned at 3 µm, and mounted on glass slides. Staining with Gill’s hematoxylin and eosin was performed according to a standard protocol. For calcium identification, alizarin red S staining (A5533-25G, Sigma-Aldrich, USA) was used: 2 g of the dye was dissolved in 100 mL of distilled water. Sections were stained for 3 minutes, dehydrated in three changes of absolute isopropanol, and mounted.

To differentiate calcified areas from cartilage, trichrome staining was applied. After deparaffinization and hydration in distilled water, slides were incubated in Weigert’s iron hematoxylin (prepared by mixing equal parts of Weigert’s Hematoxylin A and B, Labico LLC, Russia) for 5 minutes, rinsed in distilled water, and immersed for 2 seconds in 1% aqueous acetic acid. After additional rinsing, the slides were sequentially treated with 0.05% Fast Green FCF C.I. 42053 (2353-45-9, Sigma-Aldrich, USA) for 1 minute, 1% acetic acid for 30 seconds, and 0.1% Safranin O C.I. 50240 (477-73-6, Sigma-Aldrich, USA) for 10 minutes. Final dehydration was performed in absolute isopropanol (three changes), followed by xylene clearing and mounting.

### Immunostaining

Immunohistochemical staining of IVD tissue was performed to detect aggrecan, SP, NGF (all at 1:150 dilution, rabbit polyclonal antibodies, Cloud-Clone Corp., USA), and S-100 (1:150, mouse monoclonal antibodies, Cloud-Clone Corp., USA). The IHC procedure followed a manual protocol [24]. Secondary antibodies included Goat Anti-Mouse IgG H&L (AlexaFluor® 488, ab150113- 500, Abcam, USA) for S-100, Goat Anti-Rabbit IgG H&L (AlexaFluor® 594, ab150080, Abcam, USA) for NGF, and OPAL700 (from the OPAL fluorochrome series for the Mantra 2 Quantitative Pathology Imaging System, Akoya Biosciences, Marlborough, MA, USA) for SP. Cell nuclei were counterstained using Fluoroshield Mounting Medium with DAPI (ab104139-20, Abcam, USA).

### Morphometric Analysis

For light microscopy, an Olympus CX43 microscope with Olympus SC50 camera (Olympus Co., Tokyo, Japan) and ScanScope CS scanner (Leica Biosystems, Deer Park, IL, USA) were used. For immunofluorescence imaging, a ZEISS Axio Imager.Z2 microscope (Carl Zeiss Vision, Jena, Germany) was employed. Areas showing staining for all three markers in identical locations were considered artifacts and excluded from analysis. Image analysis was performed using QuPath software. Expression intensity was graded on a scale from 1 to 3.

The stage of degeneration was determined according to the Sive criteria [25], where a score of 0-3 was considered normal, 4-9 indicated degeneration, and 10-12 indicated severe degeneration.

### Statistical analysis

The Graph Prism 8.0.1 (GraphPad, La Jolla, CA, USA) was used for statistical analysis. The distributions were determined to be parametric by Shapiro-Wilk testing. All measurement data were expressed as median [Q1, Q3]. The Mann-Whitney U test was used to compare quantitative data between the two groups. A p-value < 0.05 was considered statistically significant.

## Results

### ECM degradation in the IVD

The histological degeneration score of the IVD, according to Sive’s criteria, was significantly higher in the patient group compared to healthy controls: 5.000 [3.000, 6.750] vs. 1.000 [0, 1.000], respectively (Mann-Whitney U test: P = 0.0001).

In 36.1% of patients with DDD, H&E staining revealed calcifications within the disc in the form of rarefied, dark-pink regions of rod-like or rounded morphology. In some areas, these merged into continuous zones with scalloped borders (Figure 1A). Trichrome staining also revealed calcifications as pinkish-lilac areas, similar in appearance to those seen with H&E staining (Figure 1B). Calcifications were more prevalent in the NP and often associated with fissures. Hyaline cartilage of the EPs was stained claret red (Figure 1B).

**Figure 1.**
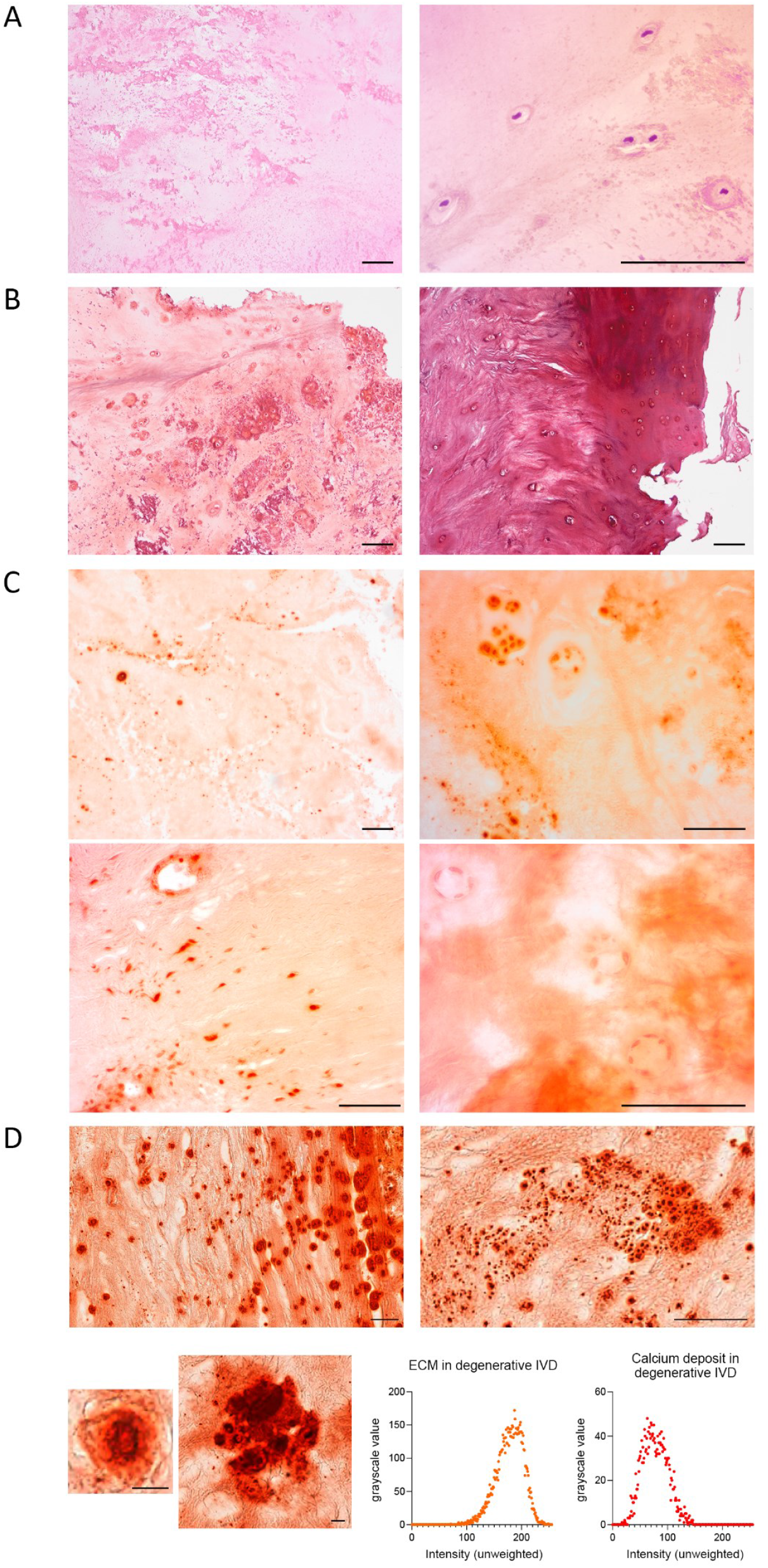
Calcification of intervertebral disc (IVD) tissue in patients with varying degrees of DDD. A. Hematoxylin-eosin staining. B. Trichrome staining: left – nucleus pulposus (NP); right – transition from annulus fibrosus to the vertebral endplate. C. Alizarin red staining: upper panel – NP; lower panel – annulus fibrosus and hypertrophied chondrocyte clusters in the NP. Scale bar for A-D: 100 µm. D. Individual calcifications in IVD tissue stained with alizarin red, imaged using a histology scanner. Scale bar for top two images: 50 µm; for bottom images: 15 µm. Comparison of grayscale intensity distribution between calcified areas and IVD extracellular matrix (ECM) using the 0-255 scale in ImageJ.

Alizarin red staining of IVD samples was irregular and nonspecific, occurring both centrally and peripherally, near hypertrophied chondrocytes, cell clusters, and intact chondrocytes. In acellular regions, a range of staining patterns was observed – from bright red to pale orange (Figure 1C). In hyaline cartilage of the endplates, darker areas likely represented denser, mineralized matrix. Chondrocyte nuclei stained darker than the ECM, but lighter than the calcifications; the cytoplasm showed the same color as the surrounding ECM. Endothelial cell and fibroblast nuclei within the AF (Figure 1C) also exhibited relatively dark staining. No artifactual peripheral or fissure-related enhancement of staining was observed.

On histological scanner images, calcifications exhibited distinct black outlines – occasionally interrupted by fissures – surrounding bright red centers of higher intensity than the adjacent disc tissue. The core of small calcifications appeared brown, occupying approximately 80% of the total area (Figure 1D). Some calcifications were polygonal, with length up to three times their width. The smallest measured approximately 1.7 µm. Notably, larger calcifications displayed various internal black striations. According to grayscale analysis, calcifications had an average intensity of 77.40 ± 22.76, compared to 178.66 ± 24.23 for the ECM.

### Aggrecan

In aggrecan-stained sections, the IVD tissue from DDD patients exhibited markedly reduced fluorescence intensity compared to control samples (Figure 2A). A significant loss of aggrecan was observed in 7 patients (19.4%; 3 females and 5 males; median age: Me = 33.00 [20.00, 36.00]), which correlated with the duration of the current episode of LBP (r = 0.449; P = 0.031; duration in weeks: Me = 6.00 [4.00, 12.00]).

**Figure 2.**
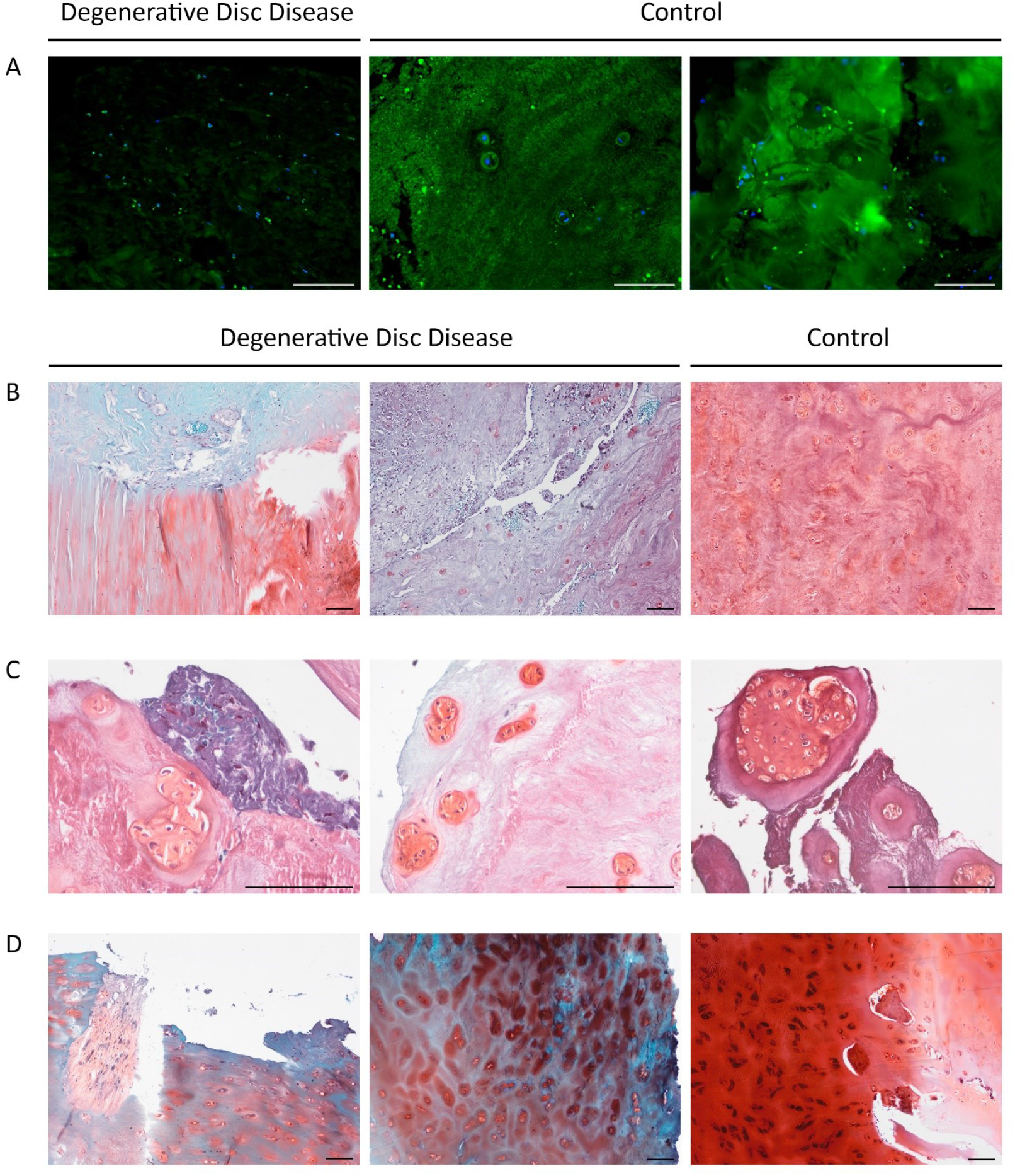
Degradation of the extracellular matrix in degenerative disc disease (DDD): reduced aggrecan content, erosive changes, and fissures. A. Aggrecan immunofluorescence in nucleus pulposus (NP) regions of DDD and control samples. B. Safranin O staining. Left: transition zone between NP and annulus fibrosus (AF). Center and right: NP region. C. Safranin O staining of degenerated NP with chondrocyte clusters. D. Vertebral endplate in DDD patient; blue staining reflects degeneration. Black-stained chondrocytes on the right may indicate pigment accumulation due to cell stress or apoptosis. Scale bar: 100 µm.

The extent of Fast Green-stained area, indicative of ECM pathology, also correlated with pain episode duration (r = 0.455; P = 0.029). Figures 2B-D show NP, AF, and endplate regions stained with Safranin O. ECM color ranged from light turquoise and pink to red-pink, violet, and blue. Chondrocyte clusters were darker and predominantly red, reflecting increased metabolic activity.

In DDD patients, the vertebral EPs exhibited erosive changes and fissures. The ECM stained intensely blue, indicating loss of hyaline cartilage components. However, chondrocytes and the narrow pericellular matrix retained a carmine hue, suggesting preserved constitutive expression. In one DDD patient, chondrocytes stained black (Figure 2D, right panel), possibly due to pigment accumulation related to cellular stress or apoptosis.

No statistically significant differences in Safranin O-stained NP tissue area were found between patient and control groups. Similarly, the two main Fast Green staining types – turquoise and blue – did not differ between groups, indicating substantial ECM heterogeneity even within individual patient cohorts.

Histological degeneration scores did not correlate with age (r = 0.138, P = 0.5), supporting the pathological rather than age-related nature of the observed degeneration.

### Expression of NGF, S-100, and SP in the IVD

The percentage of NGF-positive cells in the NP was significantly higher in DDD patients compared to controls (P = 0.0287) (Figure 3A). Notably, the highest relative expression of NGF in the control group was 7.95%, and 8 DDD patients exhibited no NGF immunoreactivity. Among the markers examined, NGF showed the strongest expression, reaching the maximum score of 3 out of 3 (Figure 3B). NGF expression was most strongly correlated with SP levels (r = 0.785; P = 5.268 × 10^−9^), suggesting their potential involvement in pain generation in DDD. This correlation may reflect a shared regulatory mechanism, such as the activation of sensory nerve endings and enhanced nociceptive signaling. In contrast, SP expression was virtually absent in control samples.

**Figure 3.**
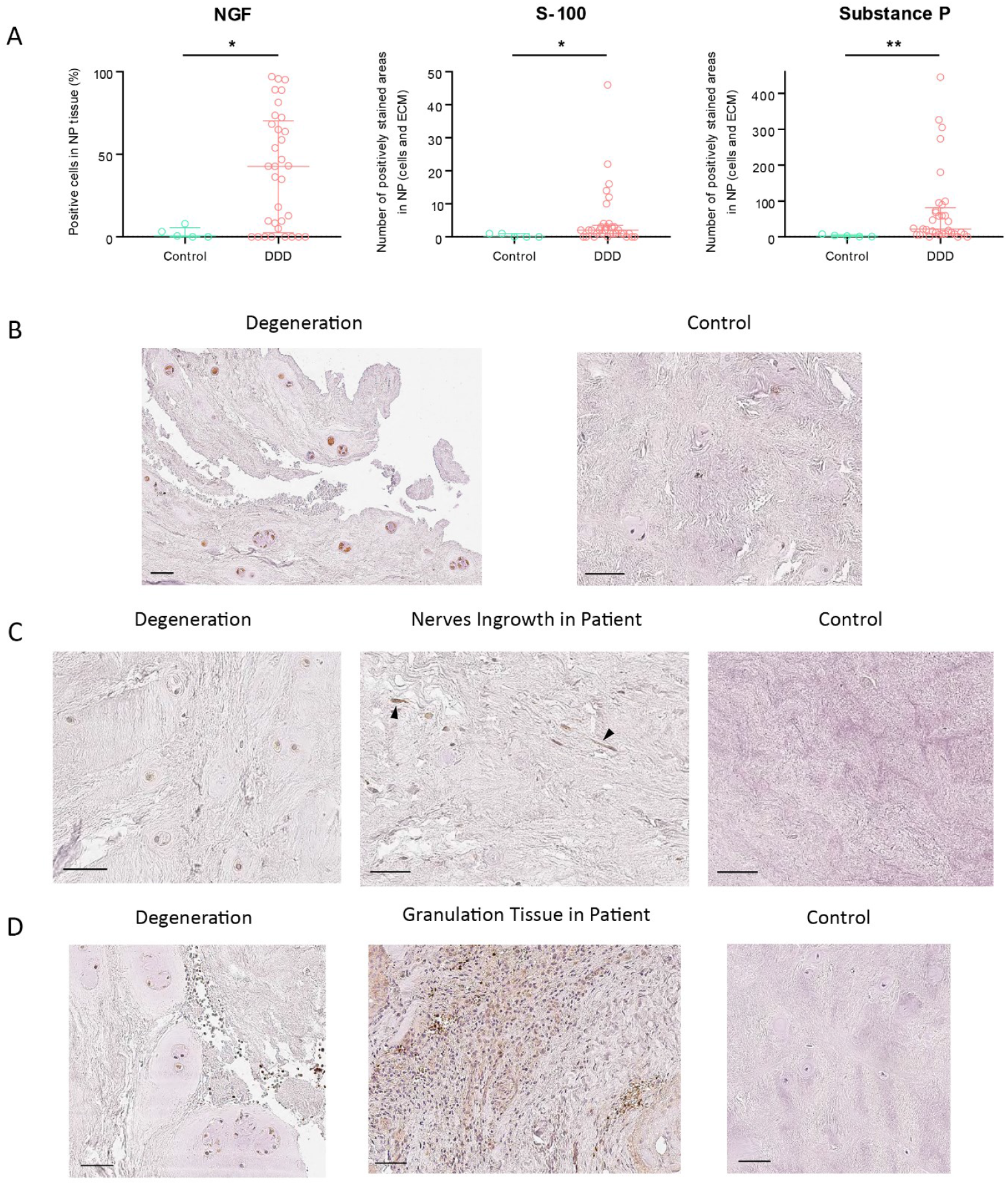
Immunohistochemical (IHC) expression of NGF, S-100, and Substance P (SP) in NP cells of degenerative disc disease (DDD) patients and healthy controls. A. Comparison by Mann-Whitney U test. B. NGF IHC. C. S-100 IHC. D. SP IHC. Scale bar: 50 µm. p < 0.05, p < 0.01. ECM – extracellular matrix, NP – nucleus pulposus.

One outlier patient presented an exceptionally high number of S-100- positive cells (n = 380) and was excluded from the analysis. S-100 expression in NP cells and the cartilage matrix was significantly higher in DDD patients compared to controls (P = 0.0312) (Figure 3B), with staining intensity scored as 1 out of 3 (Figure 3C). Free nerve endings were observed in 5 DDD patients, while none were present in controls, indicating pathological neo-innervation of the IVD.

SP expression showed the greatest differential between degenerated and healthy discs among all markers, in both NP cells and ECM (P = 0.0039) (Figure 3C), with a moderate expression level scored as 2 out of 3 (Figure 3D). Although SP expression correlated most closely with S-100 levels, the correlation was weak and statistically nonsignificant (r = 0.281; P = 0.088).

Triple immunofluorescent staining was performed to assess co-localization of the three markers. In DDD samples, a dominant violet fluorescence background from SP staining was observed, which did not overlap with the S-100 signal, thus excluding autofluorescence (Figure 4A). Autofluorescence, most prominent in the S-100 detection channel, was minimized to reveal tissue architecture.

**Figure 4.**
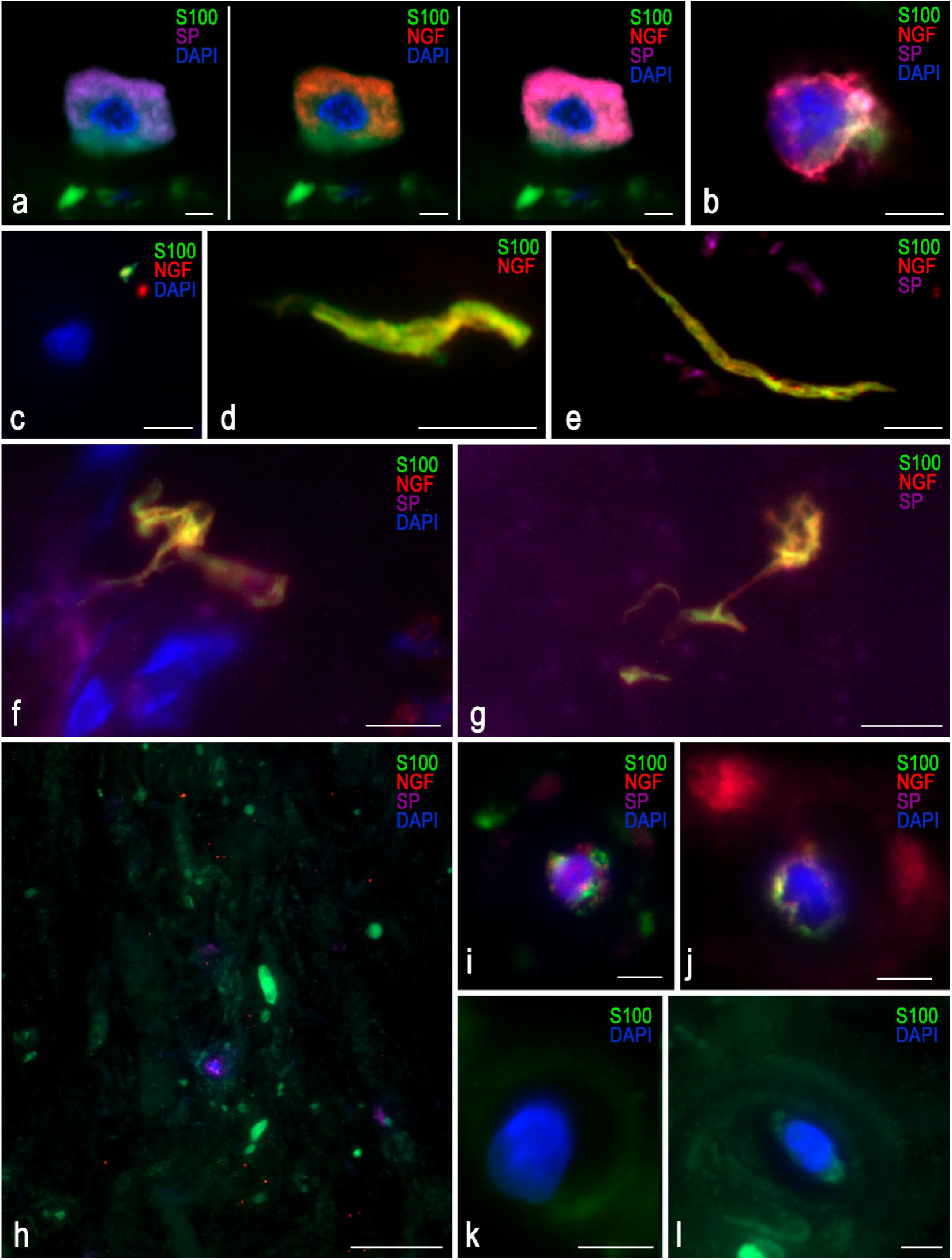
Immunofluorescent staining of intervertebral disc (IVD) tissue for S-100 (green), NGF (red), and Substance P (SP, violet). A-G: Samples from degenerative disc disease (DDD) patients. Images show a presumptive mast cell (A) and lymphocyte (B). A nerve fiber is shown growing near a chondrocyte, with NGF localized nearby; the chondrocyte’s fluorescence is restricted to its nucleus (C). A nerve fiber lacking SP expression (D). Nerve fibers in topographical proximity to SP (E-G). H-I: Control samples. Non-specific staining was observed in areas of tissue densification, particularly for S-100. Only a few chondrocytes demonstrated visible NGF and SP staining (H-J). Scale bar: 5 µm.

In IVD tissue, NGF appeared as discrete puncta up to 5 µm in diameter, including in control samples. NGF was located near S-100-positive regions and around free nerve endings, with an average distance of 0.5 µm between markers. Within an IVD fissure, a cell morphologically resembling a mast cell was observed; however, further staining for tryptase or chymase was not conducted. A presumptive lymphocyte was also identified. Figure 4B shows isolated chondrocytes with constitutive expression of all three markers.

### Correlation Analysis

We found a positive correlation between the expression levels of NGF and SP and the degree of histological disc degeneration (r = 0.467, P = 0.010 and r = 0.611, P = 0.001, respectively) (Figure 5), indicating a pathogenetic link between neuroinflammation and cartilage degeneration, independent of age. An inverse correlation between disc degeneration and age within the DDD group (r = –0.471, P = 0.015) highlights the pathological nature of the process.

**Figure 5.**
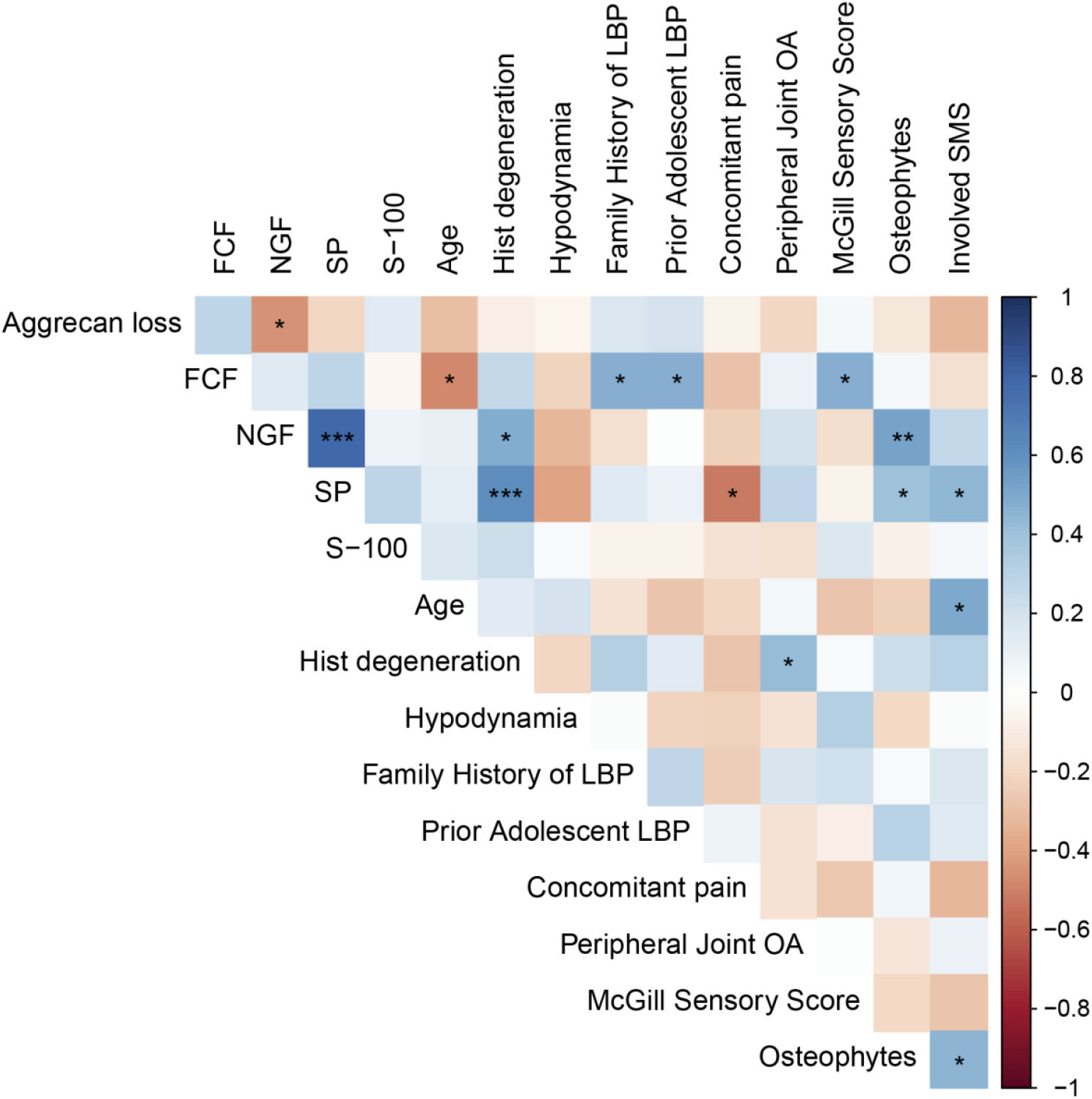
Correlation between clinical-imaging findings and histochemical/ immunohistochemical markers (Spearman’s criterion). Note: FCF – Fast Green FCF stain; SP – substance P; LBP – low back pain; OA – osteoarthritis; SMS – spinal motion segment.

Both NGF and SP expression correlated with the presence of 3–5 marginal osteophytes at the lumbar level (r = 0.526, P = 0.008 and r = 0.409, P = 0.047, respectively), supporting an association between spinal spondylosis, disc neo- innervation, and nociceptive back pain. The number of spinal motion segments affected, as determined by MRI (disc herniation combined with osteophytosis and facet joint osteoarthritis), also correlated with SP expression (r = 0.446, P = 0.029).

A history of adolescent back pain was linked to more severe adult disc degeneration with associated calcification (r = 0.476, P = 0.0216), possibly reflecting a genetic predisposition to early axial skeletal degeneration. These patients also had a positive family history of LBP (r = 0.474, P = 0.0222).

## Discussion

Aggrecan degradation is a hallmark of various degenerative conditions and has been extensively studied in osteoarthritis [26]. Rutges JP et al. proposed a grading system for NP degeneration using Safranin O staining: intense red staining (F0) decreases gradually, transitioning through a mixed red-green appearance (F1), to weak staining dominated by Fast Green (F2) [27]. In a mouse model of endplate sclerosis, 20-month-old mice exhibited dark blue Fast Green staining, in contrast to the turquoise seen in 3-month-old mice [28]. In our study, aggrecan staining in human IVD tissue appeared pink, resembling that in murine models [29]. We found no correlation between Fast Green staining intensity – indicating histological degeneration – and patient age. The relationship between aggrecan loss and calcification remains poorly understood in both animal models and human joints, including IVDs.

The presence of nerve fibers in the NP has been associated with IVD degeneration and back pain. Neo-innervation may take various forms: free nerve endings, Ruffini corpuscles, Aδ- and C-fibers, and perivascular nerves [14]. Our findings confirmed high NGF expression in NP and surrounding ECM, indicating local neo-innervation during IVD degeneration. Co-localization of NGF and SP suggests a link between nerve growth and discogenic pain. Notably, SP fluorescence was diffusely present in the ECM of degenerated discs, particularly in elongated, branching nerve fibers. Previous studies showed that anti-NGFβ treatment reduced axonal length and SP expression in NP, but had no effect on the AF [30].

Although S-100 expression in IVD chondrocytes is poorly characterized and more typical of CNS glial cells, the observed differences suggest its possible involvement in DDD pathology. The lack of correlation with S-100 expression may reflect variability in tissue sampling during surgery, as innervation – even in degenerating discs – is not uniformly distributed throughout the NP.

A relatively understudied degenerative phenotype is disc calcification, involving calcium phosphate accumulation and massive cell death in the IVD [31]. Novais EJ et al. identified a high risk of degeneration at C5-6, T6-7, and L4-5 levels; however, calcification patterns varied along the spinal column [32]. It remains unclear whether this reflects biomechanical stress or a generalized degenerative cascade. Disc calcification has been associated with systemic disorders such as chondrocalcinosis, ankylosing spondylitis, pseudogout, and hyperparathyroidism, suggesting a systemic inflammatory and metabolic basis.

MRI cannot directly detect microcalcifications, although high T1 signals may suggest fatty infiltration or calcification [33–35]. Population studies have identified disc calcification in 6% of patients with degeneration, especially in the AF of all age groups (63%) [36].

Histologically, calcified human and animal discs show increased levels of calcium, inorganic phosphate, TNAP, and collagen type X. Calcification differs by disc region: in AF and endplate, it represents heterotopic calcification mediated by pyrophosphate metabolism (PPi); in contrast, NP calcification is dystrophic, characterized by amorphous calcium crystal deposition following necrosis, independent of PPi regulation [31, 37]. The mechanisms driving calcification in DDD remain unclear and warrant further investigation. In our study, ectopic cartilage calcification was present in one-third of DDD patients.

According to Rutges JP et al., DDD is accompanied by increased expression of osteoprotegerin (OPG), collagen type X, and Runx2, with OPG most strongly correlated with degeneration in both NP and AF. These changes parallel osteoarthritic processes, including hypertrophic differentiation and calcification visible on micro-CT [38]. OPG inhibits bone resorption by binding RANKL and preventing its interaction with RANK on osteoclasts [39]. IL-1β stimulation has been shown to increase RANKL mRNA expression in a rat DDD model, but RANKL alone influences catabolic factors only in the presence of IL-1β [40]. Later work from the same group showed that IL-1β induced RANKL and OPG expression in NP and AF cells from DDD patients, though the RANKL/OPG ratio remained unchanged. RANKL and OPG mRNA levels were significantly higher in NP cells than in AF cells [41].

Mast cells are known to release various angiogenic mediators and cytokines upon activation, including VEGF, FGF-2, tryptase, chymase, IL-8, TGF-β, TNF, and NGF [42]. Moreover, mast cell-derived exosomes promote collagen synthesis and proline hydroxylation in fibroblasts, mimicking TGF-β effects and contributing to fibrotic remodeling. The presence of mast cells in NP tissue may exacerbate inflammation and ECM alterations, potentially driving degeneration, though their exact role requires further study.

## Conclusion

This study identified key molecular and cellular processes underlying the pathogenesis of IVD degeneration and associated back pain. It was demonstrated that aggrecan loss and calcification are linked to pathological innervation of the IVD. The expression of NGF and SP in degenerated discs highlights their critical role in the development of nociceptive pain. These findings emphasize the importance of early diagnosis and the development of targeted therapies aimed at modulating these pathological mechanisms. Future research should focus on elucidating the interactions between chondrocytes and the ECM to prevent further progression of disc degeneration.

## Study Limitations

In contrast to the study by Rutges JP et al. [38], which utilized Alizarin Red and Mayer’s hematoxylin staining to assess disc degeneration, a different staining protocol was employed in our study. Our method cannot be recommended as a reliable diagnostic tool for DDD in routine pathological practice, primarily because IVD specimens are commonly decalcified prior to sectioning in order to avoid artifactual fissures. Additionally, the applied method makes it difficult to accurately identify chondrocytes within the disc tissue and to assess the number and composition of cellular clusters.

## Data Availability

All data produced in the present study are available upon reasonable request to the authors.

## Author Contributions

Conceptualization, N.G.P. and N.A.S.; methodology, A.A.B., O.I.P., L.A.M. and M.A.I.; validation, A.A.B., O.I.P. and M.A.I.; formal analysis, A.V.N. and A.A.B.; investigation, N.G.P., A.V.N., A.A.B.; resources, O.I.P. and D.A.A.; data curation, N.G.P. and A.V.N.; writing—original draft preparation, N.G.P., A.V.N. and A.A.B.; writing—review and editing, N.A.S.; visualization, D.A.A., N.G.P., A.V.N. and A.A.B.; supervision, A.A.K. and V.V.T.; project administration, N.A.S.; funding acquisition, N.G.P., A.V.N. and A.A.B. All authors have read and agreed to the published version of the manuscript.

## Institutional Review Board Statement

All methods were carried out in accordance with relevant guidelines and regulations. The Ethics Committee of Pirogov Russian National Research Medical University gave ethical approval for this work (protocol No. 208 dated 17 May 2021).

## Informed Consent Statement

Written informed consent was obtained from all participants.

## Data Availability Statement

The datasets used and/or analyzed during the current study are available from the corresponding author on reasonable request.

## Acknowledgments

PhD of Health Science A. V. Aksenova (Pirogov Russian National Research Medical University), neurosurgeons of the Pirogov City Clinical Hospital No. 1, M. A. Nekrasov, V. V. Babenkov, A. N. Isayev, V. A. Smirnov, D. S. Glukhov, G. V. Gabechiya, D. B. Choriyev, A. Kh. Kozhev and neurosurgeon of the Federal Center of Brain Research and Neurotechnologies FMBA Senko I. V. for help in the research.

## References

1. Hartvigsen J, Hancock MJ, Kongsted A, et al. What low back pain is and why we need to pay attention. Lancet. 2018;391(10137):2356–2367. doi:10.1016/S0140-6736(18)30480-X

2. GBD 2021 Low Back Pain Collaborators. Global, regional, and national burden of low back pain, 1990-2020, its attributable risk factors, and projections to 2050: a systematic analysis of the Global Burden of Disease Study 2021. Lancet Rheumatol. 2023;5(6):e316–e329. Published 2023 May 22. doi:10.1016/S2665-9913(23)00098-X

3. Beyera GK, O’Brien J, Campbell S. Health-care utilisation for low back pain: a systematic review and meta-analysis of population-based observational studies. Rheumatol Int. 2019;39(10):1663–1679. doi:10.1007/s00296-019-04430-5

4. Deyo RA, Mirza SK, Turner JA, Martin BI. Overtreating chronic back pain: time to back off?. J Am Board Fam Med. 2009;22(1):62–68. doi:10.3122/jabfm.2009.01.080102

5. Peng BG. Pathophysiology, diagnosis, and treatment of discogenic low back pain. World J Orthop. 2013;4(2):42–52. Published 2013 Apr 18. doi:10.5312/wjo.v4.i2.42

6. Fardon DF, Williams AL, Dohring EJ, Murtagh FR, Gabriel Rothman SL, Sze GK. Lumbar disc nomenclature: version 2.0: Recommendations of the combined task forces of the North American Spine Society, the American Society of Spine Radiology and the American Society of Neuroradiology. Spine J. 2014;14(11):2525–2545. doi:10.1016/j.spinee.2014.04.022

7. Shapiro, I.M., Risbud, M.V. (2014). Introduction to the Structure, Function, and Comparative Anatomy of the Vertebrae and the Intervertebral Disc. In: Shapiro, I., Risbud, M. (eds) The Intervertebral Disc. Springer, Vienna. 10.1007/978-3-7091-1535-0_1

8. Newell N., Little J.P., Christou A., Adams M.A., Adam C.J., Masouros S.D. Biomechanics of the Human Intervertebral Disc: A Review of Testing Techniques and Results. J. Mech. Behav. Biomed. Mater. 2017;69:420–434. doi: 10.1016/j.jmbbm.2017.01.037

9. Tang X., Jing L., Chen J. Changes in the Molecular Phenotype of Nucleus Pulposus Cells with Intervertebral Disc Aging. PLoS ONE. 2012;7:e52020. doi: 10.1371/journal.pone.0052020

10. Richardson S.M., Ludwinski F.E., Gnanalingham K.K., Atkinson R.A., Freemont A.J., Hoyland J.A. Notochordal and Nucleus Pulposus Marker Expression is Maintained by Sub-populations of Adult Human Nucleus Pulposus Cells through Aging and Degeneration. Sci. Rep. 2017;7:1501. doi: 10.1038/s41598-017-01567-w

11. Roughley P, Martens D, Rantakokko J, Alini M, Mwale F, Antoniou J. The involvement of aggrecan polymorphism in degeneration of human intervertebral disc and articular cartilage. Eur Cell Mater. 2006;11:1–7. Published 2006 Jan 18

12. Standring S. Gray’s Anatomy: The Anatomical Basis of Clinical Practice. Churchill Livingstone Elsevier; London, UK: 2020

13. Crump KB, Alminnawi A, Bermudez-Lekerika P, et al. Cartilaginous endplates: A comprehensive review on a neglected structure in intervertebral disc research. JOR Spine. 2023;6(4):e1294. Published 2023 Oct 21. doi:10.1002/jsp2.1294

14. Groh AMR, Fournier DE, Battié MC, Séguin CA. Innervation of the Human Intervertebral Disc: A Scoping Review. Pain Med. 2021;22(6):1281–1304. doi:10.1093/pm/pnab070

15. Новикова А.в., Правдюк Н.Г., Саклакова в.С., Лоломадзе з.А., Фениксов в.М., Николаев Д.А. и др. Дегенеративная болезнь диска у молодых: цитокиновый профиль и факторы ангиогенеза. вестник РГМУ. 2021; (6): 81–9. DOI: 10.24075/vrgmu.2021.061

16. Pravdyuk NG, Novikova AV, Shostak NA, et al. Immunomorphogenesis in Degenerative Disc Disease: The Role of Proinflammatory Cytokines and Angiogenesis Factors. Biomedicines. 2023;11(8):2184. Published 2023 Aug 3. doi:10S.3390/biomedicines11082184

17. Cunha C, Silva AJ, Pereira P, Vaz R, Gonçalves RM, Barbosa MA. The inflammatory response in the regression of lumbar disc herniation. Arthritis Res Ther. 2018;20(1):251. doi:10.1186/s13075-018-1743-4

18. Risbud MV, Shapiro IM. Role of cytokines in intervertebral disc degeneration: pain and disc content. Nat Rev Rheumatol. 2014;10(1):44–56. doi:10.1038/nrrheum.2013.160

19. Binch AL, Cole AA, Breakwell LM, et al. Expression and regulation of neurotrophic and angiogenic factors during human intervertebral disc degeneration. Arthritis Res Ther. 2014;16(5):416. Published 2014 Aug 20. doi:10.1186/s13075-014-0416-1

20. He M, Pang J, Sun H, Zheng G, Lin Y, Ge W. Overexpression of TIMP3 inhibits discogenic pain by suppressing angiogenesis and the expression of substance P in nucleus pulposus. Mol Med Rep. 2020;21(3):1163–1171. doi:10.3892/mmr.2020.10922

21. Ahmed AS, Berg S, Alkass K, et al. NF-κB-Associated Pain-Related Neuropeptide Expression in Patients with Degenerative Disc Disease. Int J Mol Sci. 2019;20(3):658. Published 2019 Feb 3. doi:10.3390/ijms20030658

22. Freemont AJ, Peacock TE, Goupille P, Hoyland JA, O’Brien J, Jayson MI. Nerve ingrowth into diseased intervertebral disc in chronic back pain. Lancet. 1997;350(9072):178–181. doi:10.1016/s0140-6736(97)02135-1

23. Donato R, Cannon BR, Sorci G, et al. Functions of S100 proteins. Curr Mol Med. 2013;13(1):24–57

24. Buchwalow I, Samoilova V, Boecker W, Tiemann M. Multiple immunolabeling with antibodies from the same host species in combination with tyramide signal amplification. Acta Histochem. 2018;120(5):405–411. doi:10.1016/j.acthis.2018.05.002

25. Sive JI, Baird P, Jeziorsk M, Watkins A, Hoyland JA, Freemont AJ. Expression of chondrocyte markers by cells of normal and degenerate intervertebral discs. Mol Pathol. 2002;55(2):91–97. doi:10.1136/mp.55.2.91

26. Empere M, Wang X, Prein C, et al. Aggrecan governs intervertebral discs development by providing critical mechanical cues of the extracellular matrix. Front Bioeng Biotechnol. 2023;11:1128587. Published 2023 Mar 2. doi:10.3389/fbioe.2023.1128587

27. Rutges JP, Duit RA, Kummer JA, et al. A validated new histological classification for intervertebral disc degeneration. Osteoarthritis Cartilage. 2013;21(12):2039–2047. doi:10.1016/j.joca.2013.10.001

28. Fan Y, Zhang W, Huang X, et al. Senescent-like macrophages mediate angiogenesis for endplate sclerosis via IL-10 secretion in male mice. Nat Commun. 2024;15(1):2939. Published 2024 Apr 5. doi:10.1038/s41467-024-47317-1

29. Roughley PJ, Mort JS. The role of aggrecan in normal and osteoarthritic cartilage. J Exp Orthop. 2014;1(1):8. doi:10.1186/s40634-014-0008-7

30. Yamauchi K, Inoue G, Koshi T, et al. Nerve growth factor of cultured medium extracted from human degenerative nucleus pulposus promotes sensory nerve growth and induces substance p in vitro. Spine (Phila Pa 1976). 2009;34(21):2263–2269. doi:10.1097/BRS.0b013e3181a5521d

31. Hristova GI, Jarzem P, Ouellet JA, et al. Calcification in human intervertebral disc degeneration and scoliosis. J Orthop Res. 2011;29(12):1888–1895. doi:10.1002/jor.21456

32. Novais EJ, Narayanan R, Canseco JA, et al. A new perspective on intervertebral disc calcification-from bench to bedside. Bone Res. 2024;12(1):3. Published 2024 Jan 22. doi:10.1038/s41413-023-00307-3

33. Bangert BA, et al. Hyperintense disks on T1-weighted MR images: correlation with calcification. Radiology. 1995;195:437–443. doi: 10.1148/radiology.195.2.7724763

34. Malghem J, et al. High signal intensity of intervertebral calcified disks on T1-weighted MR images resulting from fat content. Skeletal. Radiol. 2005;34:80–6. doi: 10.1007/s00256-004-0843-1

35. Tyrrell PN, Davies AM, Evans N, Jubb RW. Signal changes in the intervertebral discs on MRI of the thoracolumbar spine in ankylosing spondylitis. Clin. Radiol. 1995;50:377–83. doi: 10.1016/S0009-9260(05)83134-4

36. Chanchairujira K, et al. Intervertebral disk calcification of the spine in an elderly population: radiographic prevalence, location, and distribution and correlation with spinal degeneration. Radiology. 2007;230:499–503. doi: 10.1148/radiol.2302011842

37. Novais EJ, et al. Comparison of inbred mouse strains shows diverse phenotypic outcomes of intervertebral disc aging. Aging Cell. 2020;19:e13148. doi: 10.1111/acel.13148

38. Rutges JP, Duit RA, Kummer JA, et al. Hypertrophic differentiation and calcification during intervertebral disc degeneration. Osteoarthritis Cartilage. 2010;18(11):1487–1495. doi:10.1016/j.joca.2010.08.006

39. Boyle WJ, Simonet WS, Lacey DL. Osteoclast differentiation and activation. Nature. 2003;423(6937):337–342. doi:10.1038/nature01658

40. Takegami N, Akeda K, Yamada J, et al. RANK/RANKL/OPG system in the intervertebral disc. Arthritis Res Ther. 2017;19(1):121. Published 2017 Jun 2. doi:10.1186/s13075-017-1332-y

41. Sano T, Akeda K, Yamada J, Takegami N, Sudo T, Sudo A. Expression of the RANK/RANKL/OPG system in the human intervertebral disc: implication for the pathogenesis of intervertebral disc degeneration. BMC Musculoskelet Disord. 2019;20(1):225. Published 2019 May 17. doi:10.1186/s12891-019-2609-x

42. Elieh-Ali-Komi D, Shafaghat F, Alipoor SD, Kazemi T, Atiakshin D, Pyatilova P, et al. Immunomodulatory Significance of Mast Cell Exosomes (MC-EXOs) in Immune Response Coordination. Clinic Rev Allerg Immunol. 2025 Feb 20;68(1):20.

